# Implementation of Male-Specific Motivational Interviewing in Malawi: An Assessment of Intervention Fidelity and Barriers to Scale-Up

**DOI:** 10.1101/2024.09.24.24314326

**Authors:** Katherine Ničev Holland, Julie Hubbard, Misheck Mphande, Isabella Robson, Khumbo Phiri, Dorina Onoya, Elijah Chikuse, Kathryn Dovel, Augustine Choko

## Abstract

**Introduction:** Treatment interruption (TI), defined as >28 days late for ART appointment, is one of the greatest challenges in controlling southern African HIV epidemics. Negative client-provider interactions remain a major reason for TI and barrier for return to care, especially for men. Motivational interviewing (MI) facilitates client-driven counseling and improves client-provider interactions by facilitating equitable, interactive counseling that helps clients understand and develop solutions for their unique needs. Fidelity of MI counseling in resource-constrained health systems is challenging.

**Methods:** We developed a male-specific MI curriculum for Malawian male TI clients. Four psychosocial counselors (PCs, a high-level Malawian counseling cadre) received a 2.5-day curriculum training and job-aid to guide MI counseling approaches. They participated in monthly phone-based discussions with their manager about MI-based solutions to challenges faced. PCs implemented the MI curriculum with men >15 years who were actively experiencing TI. Clients were found at home (through tracing) or at the facility (for those who returned to care on their own). MI counseling sessions were recorded, transcribed, translated into English, and coded in Atlas.ti v9. MI quality was assessed using a modified version of the validated Motivational Interviewing Treatment Integrity tool. The tool has two measures: 1) counts of key MI behaviors throughout the session (questions, reflections, etc.); and 2) overarching scores (using a five-point scale) that characterize three MI dimensions for an entire counseling session (cultivating change talk, partnership, and empathy).

**Results:** 44 MI sessions were recorded and analyzed between 4/1/22-8/1/22. 64% of counseling sessions focused on work and travel as the main reason for TI. 86% of sessions yielded client-driven, tailored solutions for overcoming TI. PCs implemented multiple MI behaviors very well: asking questions, giving information, simple reflections, and client affirmation. Few PCs used complex reflection, emphasized autonomy, or sought collaboration with clients. Among overarching MI dimensions, HCWs scored high in partnership (promoting client-driven discussions) and cultivating change talk (encouraging client-driven language and behavior change confidence) but scored sub-optimal in empathy. Only 5 sessions had confrontational/negative PC attitudes.

**Conclusions:** PCs implemented MI with fidelity and quality resulting in tailored, actionable plans for male re-engagement in HIV treatment in Malawi.

**Clinical Trial Number:** NCT05137210 and NCT04858243

## Introduction

Treatment interruption (TI) from HIV care is one of the most pressing challenges to controlling HIV epidemics in eastern and southern Africa [1–3]. In Malawi, TI is defined as being >28 days late for an ART refill appointment. In the region, it is estimated that 32-46% of clients on ART experience TI within the first five years on treatment [4–6], and 30-40% of clients experience repeat TI [7–8]. Men experience TI at a 15-26% higher rate than women [9] and have worse HIV outcomes like increased morbidity, mortality, and viremia [10–11]. As such, while countries like Malawi are quickly approaching aggregate UNAIDS 95-95-95 targets, young men aged 15-34 have largely fallen behind [12].

Resource limitations have been noted to increase rates of provider compassion fatigue and withdrawal [13], resulting in poor client-provider interactions that both contribute to male TI and diminish male return to care [14]. Providers often describe men as being “ill-informed,” “difficult,” “selfish,” and overall “bad clients [15].” These biases may reflect in inequitable client-provider power dynamics and can result in men avoiding HIV services [14,16]. The hierarchical structure of health systems further entrenches this by positioning providers as the expert on a client’s situation rather than as a collaborator working alongside the client [13]. The result is predominantly didactic, authoritative counseling that fails to address individualized needs of clients [13,17]. Given these issues, the World Health Organization (WHO) has increasingly called for HIV services geared toward person-centered approaches [16].

Person-centered care (PCC) is an umbrella term for services that center client respect, client engagement in informed decision-making, and support for holistic client needs (physical, psychological, and social) [14,18–20]. Specific PCC techniques, like motivational interviewing (MI), promote responsive counseling and client-driven solutions that may help to address barriers to care. MI is a counseling technique aimed at strengthening a clients’ internal motivation, commitment to behavioral change, and development of individualized action plans that promote sustained behavior change [21]. MI has been associated with increased ART adherence and decreased incidence of sexual risk taking [22,23], and qualitative data suggests client-driven counseling techniques such as MI are deeply desired by male ART clients [24,25].

Despite its advantages, MI is infrequently utilized given the extensive resources required that are rarely available in these low-resource settings (e.g., increased personnel time, higher level of personnel training, and increased monitoring and evaluation). Other challenges to regional implementation include poor fidelity (including poor uptake of “the spirit of MI” and its core behaviors, namely empathy and reflective listening) [26,27], and poor session attendance [22]. Furthermore, there is no evidence on MI as a tool for men living with HIV (MLHIV).

To improve client-provider interactions and support men experiencing TI in Malawi, we developed a male-specific MI curriculum and assessed quality and fidelity of its implementation by psychosocial counselors (PC).

## Materials & Methods

### Setting

We developed a male-specific MI curriculum for male TI clients and implemented it across 8 facilities in Malawi from April 1-August 31 2022. All health facilities were supported by Partners in Hope, a PEPFAR implementing partner that supports HIV service delivery in over 100 facilities serving >200,000 people living with HIV across Malawi. We purposively selected facilities to include large public district hospitals (n=3) and small health centers (n=5), in the central (n=4) and southern (n=4) regions.

### Male-Specific MI Curriculum

The MI curriculum was composed of two parts. First, an introductory guide was developed to describe the MI approach and help PC’s develop the needed skills to implement MI. This included guidance on how to (1) build rapport with clients, including discussing their hopes and goals for the future; (2) support clients to identify their personal barriers to care; (3) explore the potential benefits of treatment adherence and connect these benefits with the clients’ goals; (4) encourage clients’ decision making for change, and; (5) help clients set their own short- and long-term goals related to HIV treatment. Our training materials were developed using the REDI Framework and training guide curriculum developed by Engender Health to aid in client contraception choice [28]. It is one of the few examples of MI-based client-centered care in the region. Second, a male-specific MI job aid was developed to be used during one-time counseling sessions. We used literature on barriers to men’s HIV care [29–31] and formative qualitative research [15, 32–37] to identify 11 key domains related to barriers to men’s use of HIV treatment. The job aid provided an overview of each topic to sensitize the PC on common challenges to men’s use of HIV treatment and probes to understand the unique barriers, priorities, and resources of each individual client. Each domain had MI related discussion prompts to facilitate client-led discussions and solutions. These were organized by ‘Think’ and ‘Act’ questions. ‘Think’ prompts were used to help the client identify the barrier to care or to reframe the challenge to gain a better understanding of how it could be overcome. ‘Act’ prompts were meant to support the client’s decisions for change and were used to explore actionable ways of overcoming the challenge. A brief description is provided in Appendix A.

### PC Training

Four PC, a high-level Malawian counseling cadre, received a 2.5-day curriculum training comprised of 1) sensitization to the challenges and needs of men as ART clients; 2) education on best practices for male-specific MI, including building rapport, exploring barriers to HIV care, decision making for overcoming challenges, and strategizing how to implement them; and 3) practice of key MI skills through role play and peer feedback. A key focus throughout the training was the co-creation of actionable next steps for improving clients ART adherence. After the training, PC participated in monthly phone-based discussions with their manager for group feedback and discussion about challenges and solutions for implementing the male-specific MI curriculum.

### Implementation

Men who were ≥15 years, living in study facility catchment areas, and not currently in HIV care (defined as never initiated ART or experiencing treatment interruption (missed an ART appointment ≥7 days or defaulted (out of care >28 days)) or having a high viral load (>1,000 copies/ml) were offered the male-specific MI curriculum. Routine medical charts and health registers were reviewed to identify potentially eligible men. Each PC was responsible for MI implementation at two health facilities. Per standard of care, PC actively traced potentially eligible men to offer male-specific MI counseling. Men were recruited either through this community tracing, or at health facilities if they returned for their ART refill and were flagged as recently having a TI or high viral load. All identified men were screened for eligibility and, if eligible, asked for oral consent to record a one-time MI counseling session.

### Measuring Fidelity & Quality in Counseling Sessions: MITI

We measured the quality of MI components within counseling sessions using a modified version of the Motivational Interviewing Treatment Integrity tool (MITI), a validated scale to assess MI competencies and provide feedback on the fidelity and quality of MI interactions [38]. We utilized two key measures from the tool: 1) objective counts of key MI behaviors throughout the session; and 2) subjective overarching ‘global’ ratings (using a five-point Likert scale) for domains characterizing the ‘spirit’ of MI (see Appendix B). Some domains from the tool (including Softening Sustain Talk, Persuade with Permission and Persuade) were not included in our analysis due to the cultural context and the fact that some themes from MITI cannot be measured well via transcripts. Behaviors were classified as “simple” versus “complex” in accordance with whether the behavior in question is common to traditional counseling (e.g., asking questions, giving information, simple reflection, affirmation) versus if the behavior is more unique to MI-based counseling (e.g., complex reflection, seeking collaboration, emphasizing autonomy).

In addition to the MITI tool measures, we also assessed PC’s abilities to problem solve and develop next steps to overcome adherence challenges. We rated sessions as ‘effective’ if actionable next steps were created to address the session’s client-identified barriers, or ‘not effective’ if no actionable next steps were created.

### Data Analysis

MI counseling sessions were recorded, transcribed, and translated into English for analysis. We completed structured memos based on the MITI scale to measure fidelity and quality, including frequency of MI behaviors and ratings of MI domains. Transcripts were reviewed and key data was extracted and entered into the memo, including simple excerpts of text that reflected strong or weak implementation of MITI domains. Transcripts were simultaneously coded in Atlas.ti v.9 using codes for each of the MITI behaviors to aid in the behavior counts and domains to ensure all content related to themes were captured in their entirety. Client reasons for TI, counts of global rating scores and key MI behaviors per interview were analyzed in Excel.

### Ethics Statement

The IDEaL trial is registered with ClinicalTrials.gov (NCT05137210) and was approved by the UCLA and NHSRC of Malawi IRB. Men were recruited either through this community tracing, or at health facilities if they returned for their ART refill and were flagged as recently having a TI or high viral load. All identified men were screened for eligibility and, if eligible, asked for oral consent to record a one-time MI counseling session.

## Results

Overall, 44 counseling sessions were conducted. Counseling session duration ranged from 6-60 minutes, with an average of 27 minutes (IQR: 18.75). All sessions were completed by male PCs with a minimum of three years’ counseling experience prior to the study (mean: 5.65 years’ experience).

### Client Type and Reason for TI

Client type was recorded for 36/44 counseling sessions. Twenty men (45%) had been out of care for > 28 days, fifteen (34%) had missed an ART appointment by 7 days, one (2%) had high viral load, and 8 (18%) were unrecorded. Across the 44 counseling sessions, participating men most frequently cited “work & travel” (n=28, 64%) as their main reason for TI. Other reasons for TI included difficulty “remembering appointment and medication schedules” (n=5, 11%) and feelings of “loneliness, isolation, and lack of social support” (n=4, 9%).

### Application of Key MI Behaviors

We measured frequency of the eight key MI behaviors throughout each counseling session. While no ‘target frequency’ exists for any of the MI behaviors, PC most frequently employed simple MI behaviors (e.g., giving information, asking questions, simple reflection, and affirmation). Advanced behaviors (e.g., complex reflection, seeking collaboration, and emphasizing autonomy) were less frequently utilized. PC overwhelmingly used the seven positive MI behaviors and rarely used the one negative behavior, confrontation, marked as being antithetical to the ‘spirit of MI.’ Only 11% of counseling sessions (n=5) had one or more instances where the PC demonstrated confrontational behavior (Table 1).

**Table 1.** Key MI Behavior Instances. Mean number of behavior instances per interview, with variance

| MI Behavior | Mean | Standard Deviation |
| --- | --- | --- |
| Giving Information | 8.32 | 7.40 |
| Asking Questions | 35.45 | 26.09 |
| Simple Reflection | 5.18 | 5.20 |
| Complex Reflection | 0.80 | 1.13 |
| Affirmation | 4.07 | 3.56 |
| Seeking Collaboration | 0.36 | 0.69 |
| Emphasizing Autonomy | 0.32 | 0.77 |
| Confrontation (Negative Behavior) | 0.16 | 0.53 |

### Most Frequently Used Behaviors

PC relied heavily on the four simple MI behaviors. Giving information and asking questions were the most frequently utilized. Information was given to resolve knowledge gaps while asking questions worked to identify these gaps and push the conversation deeper. PC utilized simple reflection primarily to help clients feel heard and understood. PC echoed client’s words back to them to indicate that they were paying attention or to give the client opportunity to clarify and reinforce their own thought processes. PC used affirmation to acknowledge a client’s positive attributes and to encourage the positive aspects of a client’s behavior even when the overarching behavioral theme resulted in negative outcomes, like treatment interruption (Table 2).

**Table 2.**
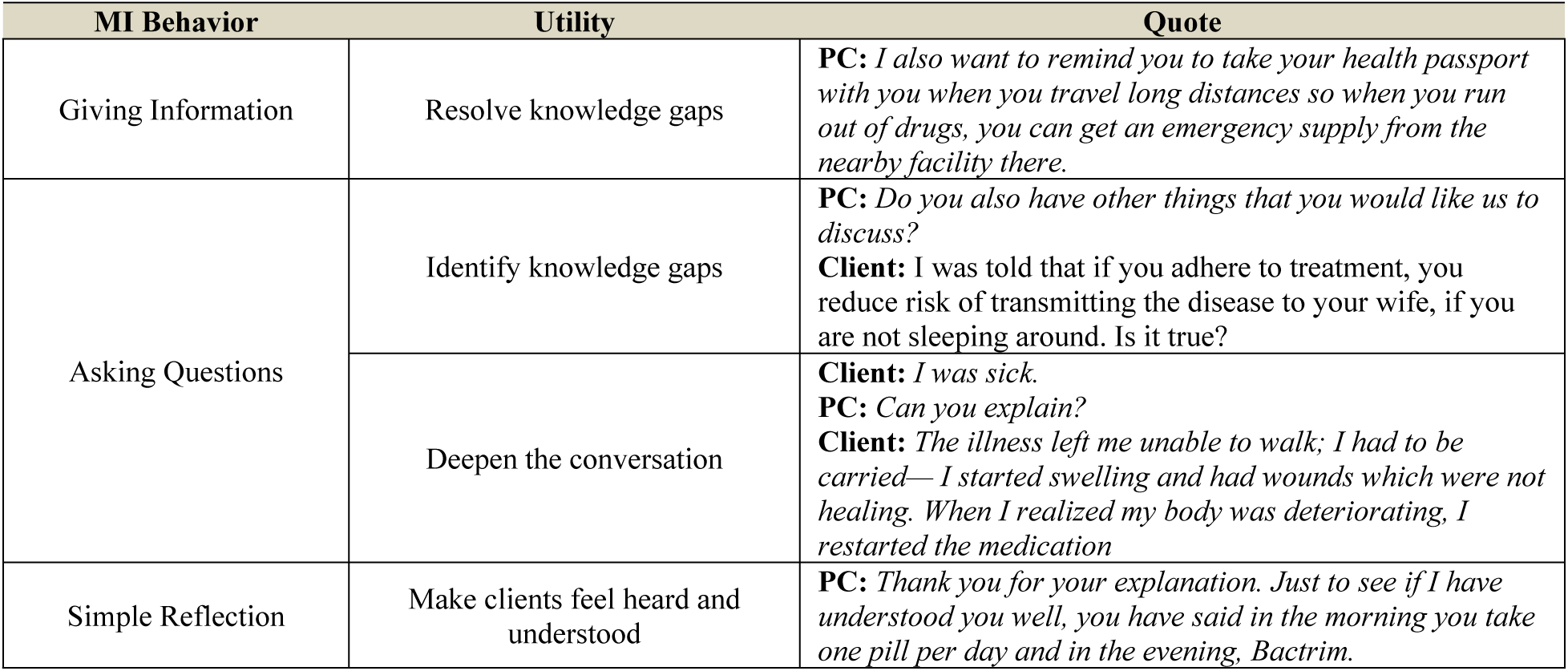
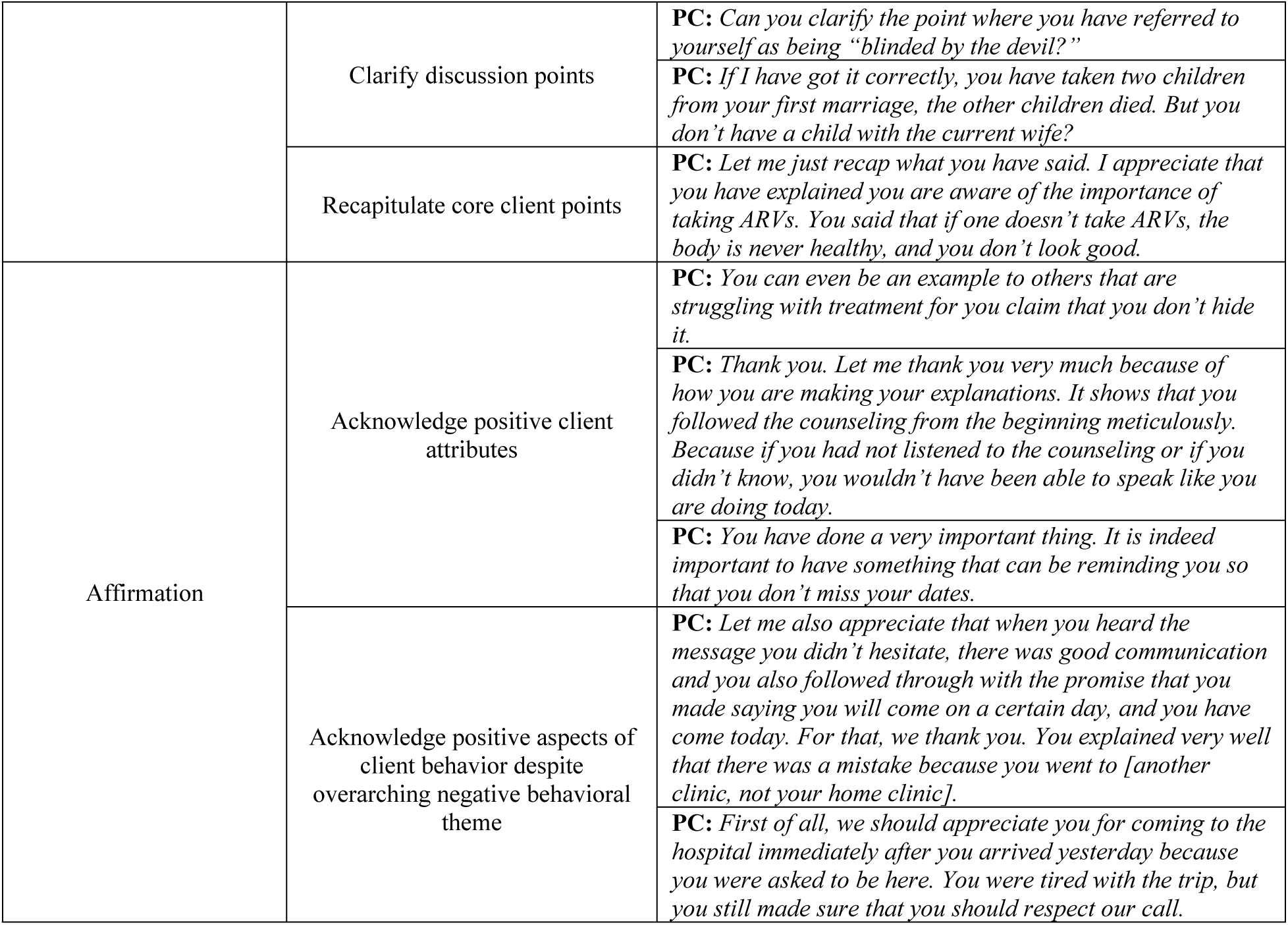
Qualitative Examples of Frequently Utilized MI Behaviors in Action. Summary of most used MI behaviors, with example quotes

### Least Frequently Used Behaviors

PC rarely used advanced MI behaviors. When used, complex reflections allowed PC to distill a client’s thought into key points, pushing the conversation forward by both extracting and emphasizing salient emotions and themes that arose throughout the client’s words. Even rarer was the use of seeking collaboration. However, when used, it emphasized power sharing and allowed the client to drive the conversation in a direction most suitable for their personal needs. Emphasizing autonomy was the least utilized positive MI behavior among PC. Despite this, its implementation was critical to fostering client buy-in by dispelling notions of being forced or shamed into regular medication adherence.

While infrequent, confrontation (negative MI behavior) did occur. PC reverted to confrontational language when they felt their clients were not being honest and when they felt they needed to correct behaviors they viewed as negative (Table 3).

**Table 3.**
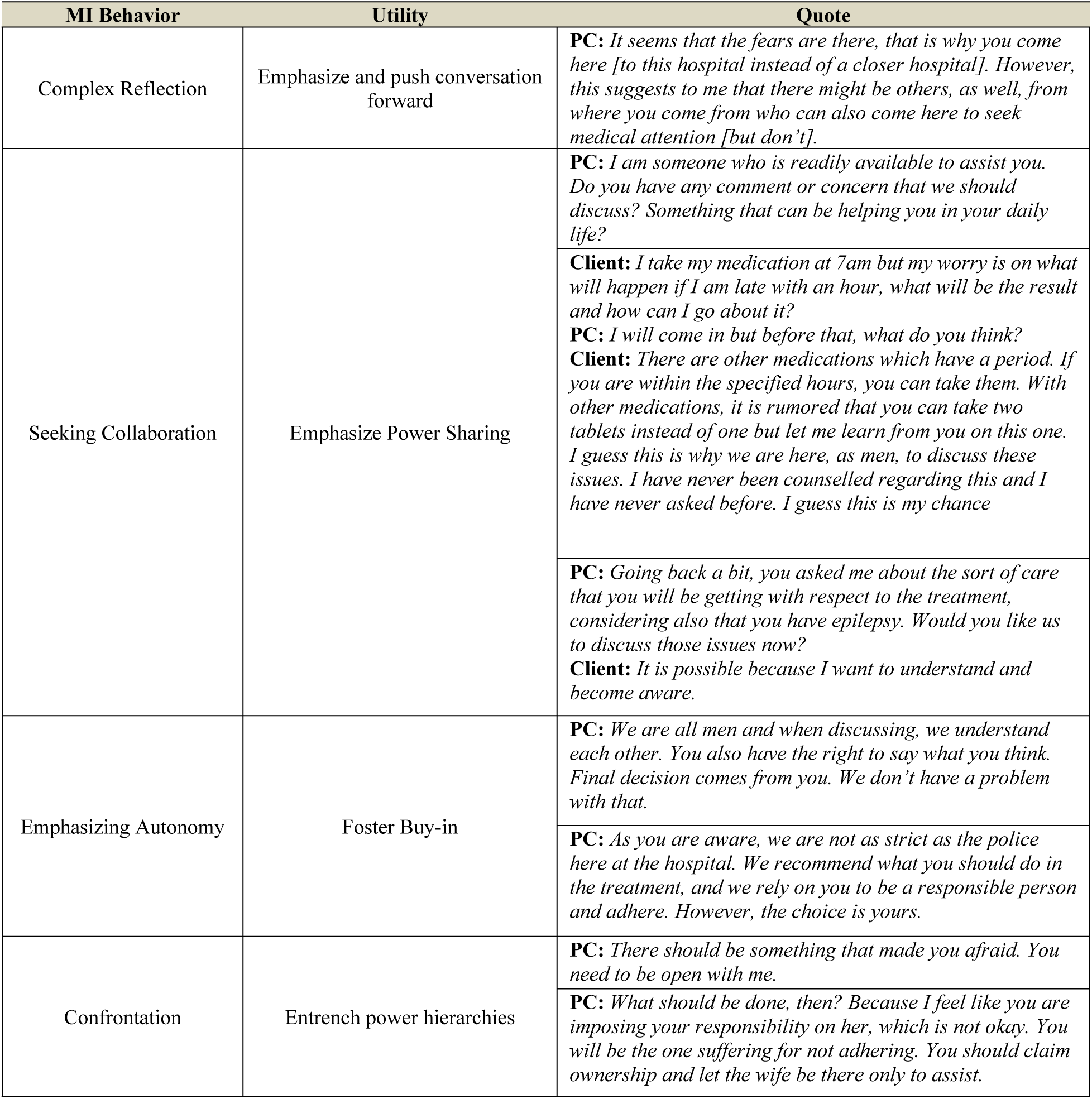
Qualitative Examples of Infrequently Utilized MI Behaviors in Action. Summary of least used MI behaviors, with example quotes

### Creating Actionable Next Steps

Across the 44 counseling sessions, ninety-one percent of counseling sessions resulted in actionable next steps for at least one barrier to care and 86% developed solutions for all identified barriers (See Appendix C examples). With the support of PC, men developed plans for overcoming barriers to care including disclosure to close male friends, posting ARV reminders on their bedroom walls, and having a plan in place for unanticipated travel, among others.

Four out of the 44 sessions did not yield actionable steps; the sessions identified as failing to yield next steps were all focused on ‘work and travel’ as the primary barrier to care. Of the four instances where no next steps were given for the topic of “work & travel,” two involved the client not having adequate transport to attend the facility and two involved one-off instances of illness while traveling which prevented return to the facility. Counseling discussions suggested particular difficulty overcoming time and cost associations with work and travel.

### Quality of MI by Global Ratings

Global ratings for elements that represented the ‘spirit of MI’ – cultivating change talk, partnership, and empathy – were graded on a 1 (lowest) to 5 (highest) scale for each counseling session (Table 4).

**Table 4.**
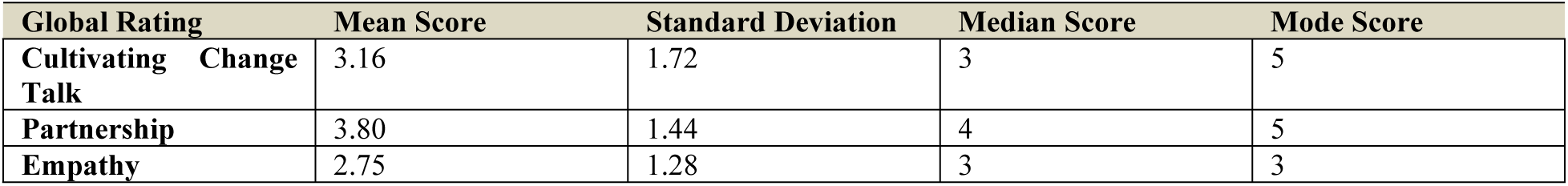
Central Tendency and Spread of Global Rating Scores Per Interview. Mean, median, and mode global rating score per interview, with variance

| Global Rating | Mean Score | Standard Deviation | Median Score | Mode Score |
| --- | --- | --- | --- | --- |
| Cultivating Change Talk | 3.16 | 1.72 | 3 | 5 |
| Partnership | 3.80 | 1.44 | 4 | 5 |
| Empathy | 2.75 | 1.28 | 3 | 3 |

### Partnership

PC generally scored best in partnership, or their ability to convey an understanding that expertise and wisdom about change resides mostly within the client, with a mean score of 3.8. PC displayed partnership through statements that highlighted wisdom residing in the client themselves and by using terminology that offered the client an equal position in the discussion. When PC used language to convey partnership, it elicited higher-level engagement from the client; rather than simply listening, the client began to actively participate and offer their critical thoughts on the subject at hand. This helped build rapport and the client’s sense of agency in crafting a plan toward treatment re-initiation. Most commonly, partnership came in the form of questions that evoked solutions from the client, but it occasionally came in the form of explicit statements calling for equality.

Despite scoring well in partnership, several exchanges between clients and PC signaled that clients were resistant to, or initially did not accept, the PC’s bid for partnership. One such example is seen below where the client appears to be either expecting or asking for a more traditional, didactic interaction with the PC:

**PC:** The medication is not compatible with you. Okay, so do you know how we solve this problem?

**Client:** *You tell me*.

### Cultivating Change Talk

PC also achieved good scores for cultivating change talk, marked by the mentor actively encouraging the client’s own language in favor of the change goal, with a mean score of 3.16.

Cultivating change talk occurred through extended back and forth dialogue between the PC and client whereby the PC would help identify a cause of treatment interruption and explore the barrier with broad, open-ended, and evocative questions that required the client to consider the issue at hand from multiple angles. The result was often that the client would come to a conclusion that aligns with traditional PC recommendations but on their own accord.

### Empathy

PC received the lowest scores for empathy, with an average mean score of 2.75. When employed, PC used empathic language to demonstrate that they saw the client as a human and understood that perfection is not always a reasonable goal. They emphasized that one must give themself grace while on treatment— the client should not wait to start treatment until they can adhere perfectly; everyone forgets at times and persevering is what matters. PC also expressed empathy by apologizing on behalf of the health system for negative experiences the client endured in the past even when the PC was not directly responsible for the negative interaction. Mostly commonly, empathy was expressed in the form of “as men” statements which sought to express to the client that the PC understood, as a fellow man, that there are many conflicting responsibilities that can make adhering to treatment particularly difficult (Table 5).

**Table 5.** Qualitative Examples of Global Rating Dimensions. Summary of global ratings, with example quotes

| Global Rating | Utility | Quote |
| --- | --- | --- |
| Partnership | Evoke client-driven solutions | **PC:** *You said you started taking ARVs a few years ago. In your own opinion, how would ARV adherence help with your other responsibilities in life?*<br>**Client:** *It would give me longer life and allow me to both care for my children and carry out the responsibilities I was elected for in my village.* |
|  |  | **PC:** *What have you learned from this situation? How can we solve this problem [of ART adherence]?*<br>**Client:** *Firstly, I have indeed missed the full month. In past times, we used to leave our phone numbers here and they would either call or visit us as a reminder, often offering ART adherence counseling, as well.* |
|  | Explicitly call for equality | **PC:** *You call me [HCW's first name]. Don't call me boss but take me as a fellow man who wants to chat and discuss two or three things with you. You will learn something from me, and I will learn something from you, too.* |
|  |  | **PC:** *What should we do so that whenever such trips happen you shouldn't compromise your treatment? I just want us to understand what prevented you from coming for refill so that together we can sort out and prevent it from happening again next time.* |
| Cultivating Change Talk | Facilitate client-driven language in favor of change goal | **PC:** *Based on what you shared, you missed your appointment because you were sharing medications with your wife, correct?*<br>**Client:** *Yes.*<br>**PC:** *When your wife is given a 3-month refill and you share with her, what happens?*<br>**Client:** *From my experience, it causes a huge problem for us and the healthcare workers because we end up missing appointments, and it has consequences.*<br>**PC:** *Do you think sharing medication is a good decision?*<br>**Client:** *It is not a good because when we are given appointments, the healthcare workers calculate how much medication we need for that period. When we share medicine, one person will not have enough.*<br>**PC:** *What should be done then? As you said, sharing medicine with your wife disturbs both of your schedules.*<br>**Client:** *I should talk with my wife and have her return to the facility since her dates and medications no longer align.* |
| Empathy | Emphasize Grace on Treatment | **PC:** *It is bound to happen because people are not computers. [They] happen to forget.* |
|  |  | **PC:** *We are all humans, and by that, we do forget things. That is normal.* |

## Discussion

We assessed quality and fidelity of male-specific MI curriculum implementation by PC in routine HIV care settings in Malawi and found that PC were able to achieve both quality and fidelity in certain areas. Following a brief training, PC could (1) effectively use most MI behaviors with rare instances of confrontation, (2) demonstrate successful application of MI domains, and (3) guide counseling sessions that resulted in actionable next steps for clients to overcome TI. However, PC demonstrated infrequent usage of advanced MI behaviors and did not display high rates of empathy with clients suggesting that additional training or altered curricula may be needed to promote advanced MI behaviors and global skills.

When implementing male-specific MI, PC in our study overwhelmingly relied on simple behaviors (asking questions, giving information, simple reflection, and affirmation). Despite their simplicity, these simple MI behaviors seemed to engage male clients by clearing knowledge gaps, making clients feel seen and heard, and affirming clients. Advanced MI behaviors (complex reflection, seeking collaboration, and emphasizing autonomy) were used less frequently but served to deepen conversations, center counseling on the unique needs of the client, and reaffirm that enduring solutions reside within the client, not the PC. Infrequent usage of advanced MI behaviors may reflect ingrained client-provider power dynamics and hierarchies that position providers as experts meant to direct, rather than collaborate, with a client [13]. Infrequent usage of complex behaviors may also reflect the need for additional training to advance the nuanced counseling skills necessary for deeper MI implementation. PCs’ ability to engage in effective MI despite minimal use of complex MI behaviors suggests that simple, paired down MI behaviors may be sufficient to instill the ‘spirit of MI’ into a counseling session in contexts such as Malawi. While infrequent, rare confrontational exchanges worked to entrench typical counseling hierarchies and fortify the unequal power dynamics that MI seeks to dismantle, thus creating a more hostile environment not conducive of open dialogue.

Despite most interviews resulting in actionable next steps and being rated as ‘effective,’ there was wide fluctuation in behavior frequency across interviews, suggesting that there is no ‘right number’ of behavior frequencies required to enact high-quality MI. Frequencies of usage should be expected to be variable, fitting with the spirit of MI, which seeks to provide an individualized approach rather than a ‘one-size-fits-all’ approach. Effective counseling sessions also varied considerably in length, further supporting that there is no one-size-fits-all approach to quality, effective MI.

Forming actionable next step plans was critical to enacting effective MI. Ninety-one percent of counseling sessions resulted in actionable next steps for at least one barrier to care and 86% developed solutions for all identified barriers. With the support of PC, men developed plans for overcoming barriers to care including disclosure to close male friends, posting ARV reminders on their bedroom walls, and having a plan in place for unanticipated travel, among others. While rare, some sessions failed to develop plans for overcoming; this overwhelmingly occurred when the reason for TI was “work & travel,” which was also the main identified reason for TI among study participants. Counseling sessions suggested that time and cost associations with work and travel may be particularly challenging to address due to their logistical nature, particularly in low-resource settings. Other literature has similarly found that many clients experience HIV treatment interruption in part due to insufficient affordable transport solutions and conflicting financial demands which are not easily resolved given current infrastructure and socioeconomic realities [39–41]. Additional novel structural interventions at the health facility level, like home ARV delivery or multi-month dispensing, may be needed to overcome these barriers, if feasible in the local context.

PC were largely proficient in two of the three core elements that represent the ‘spirit of MI,’ partnership and cultivating change talk, after only 2.5 days of training. Despite success in those domains, PC scored worse in empathy. This may reflect several challenges including, but not limited to: (1) cultural practice of expressing empathy in less explicit or non-western manners [42], (2) PC practice of expressing empathy through body language rather than words, thus not flagging in transcripts, or (3) insufficient training in engaging MLHIV with empathetic statements.

The overwhelming proportion of study counseling sessions developed actionable next steps and were ultimately rated as ‘effective,’ suggesting that brief trainings focused on developing MI behaviors and low-cost job aids which remind PC of basic MI principles may be effective tools to aid basic MI implementation in routine settings. As HIV programming transitions back to ministries of health from PEPFAR [43–45], these results bode well for continued implementation; it is possible to provide high-quality, skilled MI counseling with little resources and time with minimal upfront investment.

## Conclusion

Our findings are critical for understanding feasibility of providing high-quality MI counselling in resource-limited settings and will only become more relevant as funding for HIV programming across the continent becomes more constrained. Our results suggest that PC can implement high-quality MI with fidelity in a low-resource setting with brief training. While largely successful, additional training or altered curricula may help promote the lesser-seen behaviors and global skills, like complex reflection or empathy. Further, “work & travel” remains a chief barrier to treatment among MLHIV and a particularly challenging barrier to care for PC and clients, alike, to overcome. Future work should focus on taking male-specific MI counseling to scale in the standard-of-care through continually simplified trainings, curricula, and monitoring and evaluation strategies. Additional areas of future focus include exploring challenges to PC engagement with advanced MI skills, like complex reflection and empathy.

## Data Availability

The data that support the findings of this study are openly available in figshare at https://figshare.com/s/430c414b7401b8e22067 DOI: 10.6084/m9.figshare.26240885

https://10.6084/m9.figshare.26240885

## Authors’ Contributions

KD is responsible for funding acquisition. JH, MM, IR, EC and KD developed the male-specific counseling curriculum and study implementation protocol. JH, MM and IR implemented the study and collected all data. KNH, JH and KD contributed to the analysis conception. KNH and JH coded and analyzed the data with support from KD. KNH wrote the first draft and JH, MM, IR, KP, DO, EC, KD and AC edited following drafts. All authors have read and approved the final manuscript.

## Acknowledgements

We are grateful to the male clients and HCWs who participated in the study and our IDI enumerator, McDaphton Bellos, and FGD leads Misheck Mphande and Eric Lungu who conducted the interviews. We would also like to acknowledge Partners in Hope Malawi and thank them for their partnership and collaboration.

## Disclaimers

None

## Data Availability Statement

The data that support the findings of this study are openly available in figshare at https://figshare.com/s/430c414b7401b8e22067 ; DOI: 10.6084/m9.figshare.26240885

## List of Abbreviations

IRB: International Review Board
MI: motivational interviewing
MITI: Motivational Interviewing Treatment Integrity Tool
MLHIV: men living with HIV
NHSRC: National Health Sciences Research Council
PC: psychosocial counselors
PCC: person-centered care
TI: treatment interruption
UCLA: University of California, Los Angeles
WHO: World Health Organization

## Appendix A Topics & Descriptions in the Male Specific MI Curriculum

1 Appendix A: Topics & Descriptions in the Male Specific MI Curriculum
| Domain | “Think” Prompt | “Act” Prompt |
| --- | --- | --- |
| **Stigma & Discrimination** | Remember, HIV is a virus that affects people worldwide. There are others who have the same fears as you. You are not alone. | Do you know anyone else who is living with HIV? Let’s make a plan for you to ask to talk to them about their experience with HIV stigma and how they manage it. |
| **Status Disclosure** | Do you know anyone who has disclosed successfully? If so, how did they do it? How did they overcome their fears? If they disclosed to you, how did it make you feel? | Think about someone you want to disclose your HIV positive status to. Now let’s role play this scenario together. |
| **Disbelief & Challenges with Status Acceptance** | There are many people out there who choose not to test. You are being a responsible person who is choosing to know your status. | If you are struggling to accept your status, it may be helpful to get re-tested. This is always an option – would you like to make a plan ask the provider to re-test? |
| **Poor Knowledge of ART** | Think about things about ART that you want to understand. Do you want more information about how to protect a sexual partner? Or are you curious about how you can have children now that you are HIV positive? | Let’s go through your list of questions together and I will answer any of your questions I am able to answer. If I don’t have the answer, we can go to the provider together to ask so we both are able to learn. |
| **Side Effects** | Many medications, such as medication for bacterial infections, Malaria, TB, cold and flu, have a risk of some side effects as our bodies adjust. IARVs can be similar. How have you overcome other side effects in the past? | Some side effects can be overcome with other medications that your provider can provide (exp: anti-nausea medication). Let’s make a plan to ask your provider what medications are available to help with side effects. |
| **Loneliness, Isolation, & Lack of Social Support** | It can be good to realize you are not alone, that these feelings are human regardless of HIV status. Do you know anyone else, HIV positive or not, who has experienced loneliness or a lack of support? | This week, let’s make a plan for you to talk to one of your trusted friends about how they can help you feel supported. |
| **Taking ART when Feeling Healthy** | We often think that ART is only for sick people. But this is not true! ART is for anyone who is HIV positive, regardless of whether they are sick or well. | Would you like to start ART today? |
| **Lifelong Medication** | The grasshopper can travel long distances even with small jumps. Every day is a small step. Being on treatment for life may feel long, but taken bit by bit, it is manageable. | Taking medication everyday can be less burdensome when it’s integrated into your daily habits. What time is best for you to take your medication? Let’s link that time to a habit that you do every day (brushing your teeth in the morning, taking tea, etc). |
| **HIV, Work, & Travel** | We as men need to juggle many responsibilities at once. We can manage work commitments while still managing responsibilities like going to church, being a leader in the community, taking care of our families. Can you describe the commitments you have and how you keep these commitments and work at the moment? | What do you do if you need to take your medication at work but don’t want others to see it? Wrap the pills that you need in a small cloth pouch or plastic zip bag and keep the pills in your pocket or a safe location (i.e. a backpack) while you are away from home. |
| **Lifestyle (alcohol, etc)** | We know we can’t change everything about our behaviors at once. That’s okay. But we can make small changes every day to ensure we are living the as healthy as we can. | Think about small changes that you can make to your lifestyle to take a step towards better health (i.e. one less drink/pack of cigarettes per week, making a conscious decision to be prepared with condoms before going out, etc). What’s your change? |
| **HIV clinics** | Most health programs focus on children or mothers. But YOUR health as a man is equally important because it is the main vehicle towards reaching your goals in life and keeping your family well. Think about yourself. Do you prioritise your health? How do you do this? | Next time you're at the clinic for routine health for yourself or for someone else, consider starting ART. You are already there – you've taken the time, effort, and spent the transport money to attend. You can make the trip work double for you by deciding to start during a visit you are already at the clinic. |

## Appendix B Domains Included in the MITI Scale & Definitions

1 Appendix B: Domains Included in the MITI Scale & Definitions
| Behavior Count | Definition | Example |
| --- | --- | --- |
| **Giving information** | Mentor gives information, educates, provides feedback, or expresses a professional opinion without persuading, advising, or warning. Tone is usually neutral. | “It is recommended that you take your medication at the same time every day.” |
| **Asking Questions** | Any question that is asked. Includes fact-finding, close-ended, and open-ended questions that cannot be answered with yes or no and are phrased in a way that require further explanation from the client. | “What challenges are you facing when it comes to taking your medications?” |
| **Simple Reflection** | Instances where the mentor reflective listening statements in response to client statements. Simple reflections typically convey understanding or facilitate client–mentor exchanges. These reflections add little or no meaning (or emphasis) to what clients have said. | “You seem discouraged.”<br>“Things seem very stressful for you.” |
| **Complex Reflection** | Instances where the mentor uses reflective listening statements in response to client statements. Complex reflections typically add substantial meaning or emphasis to what the client has said. These reflections serve the purpose of conveying a deeper or more complex picture of what the client has said. | “It seems like it’s really hard for you to take medication every day.” |
| **Affirmation** | Instances that accentuate something positive about the client. To be considered an Affirm, the instance must be about client’s strengths, efforts, intentions, or worth. | “It sounds like you have worked really hard to stay adherent when traveling.” |
| **Seeking Collaboration** | Instances where the mentor explicitly attempts to share power or acknowledge the expertise of the client. The key here is that the mentor genuinely <u>seeks consensus</u> with the client regarding tasks, goals, or directions of the session. | “Since you have not disclosed your status, would it be alright if we spent some time discussing your relationship with your family/those close to you?” |
| **Emphasizing Autonomy** | Instances where the mentor clearly focuses the responsibility with the client for decisions about and actions pertaining to change. They highlight clients’ sense of control, freedom of choice, personal autonomy, or ability or obligation to decide about their attitudes and actions. | “You are the only one who can decide which direction your life will go” “At the end of the day, it’s up to you to take your medication every day.” |
| **Confrontation (Negative Interaction)** | Mentor directly and unambiguously disagrees, argues, corrects, shames, blames, criticizes, labels, warns, moralizes, ridicules, or questions the client’s honesty. Such interactions will have the quality of uneven power sharing, accompanied by disapproval or negativity. | “You said you drink multiple times during the week. This is way too much alcohol. That is not good.” |

## Appendix C Next Step Action Plans

1 Appendix C: Next Step Action Plans
| Reason for Treatment Interruption | Number of Clients with a Particular Reason for Interruption | Number of Instances with “No Next Steps” Established | Example of an Established “Next Step” for Mitigating Future Treatment Interruption |
| --- | --- | --- | --- |
| Fear of Disclosure | 1 | 0 | Client is an elder and a chief and this status puts him in a precarious position when it comes to waiting in a public line for ARVs → Clinician recommends using a guardian to obtain medications. |
| Side Effects | 2 | 0 | Client states they previously had bad reactions to ARVs → Clinician informs client that there are new medications with lower side effect profiles. Clinician also informs client that, in the event of a medical issue like medication side effects, client should call the hospital so they can be seen by a doctor. |
| Loneliness, Isolation, & Lack of Social Support | 4 | 1 | Client’s second wife recently passed away. He is not on good terms with his first wife. Further, his biological children are far from him and his stepson in an Evangelical Christian who does not believe in hospitals and thus won’t go to them. Client feels lonely in his diagnosis → Clinician suggests that client reach out and disclose to a male friend or male relative who may be able to act as a guardian. Client thinks this is feasible and plans to do so. |
| “But I Feel Healthy” | 1 | 0 | Client reaffirms that he feels healthy and is not in need of medications → Clinician uses a metaphor about a house that looks nice on the outside but is being destroyed by termites on the inside to express that even if you look or feel healthy now, your body is sick. Client says they understand now, and they will start taking medications so that “things should go smoothly.” |
| HIV & Other Responsibilities: Work & Travel | 28 | 4 | Client missed their appointment because they had to return to their childhood home to help their sick mother cultivate the maize → Clinician reminds client that, in the future, when going away from their current home they can use several methods to obtain ARVs: (1) take a health passport and collect ARVs at a hospital near where they are staying, (2) use a guardian to gather medications, or (3) upon gaining trust of a provider, consider asking for multi-month medication dispensing |
| HIV Clinics | 3 | 1 | Client states that clinic lines are too long, and they don’t have time to wait → Clinician informs client that clinic is typically much quieter after 12pm and simply arriving to clinic later in the day could resolve the client’s concerns.<br>Client expresses immense fear of the HIV clinics and explains that his friend was once screamed at for not properly adhering to his ARVs → Clinician apologizes for the friend’s experience and lets the client know that each clinic has a suggestion box available if someone has a poor experience. Clinician then walks the client through avenues the client can take to express their feelings to a clinic manager. |
| Sharing Medications | 3 | 0 | Client did not come to clinic because his wife also have HIV and he used some of her medications rather than picking up a refill → Clinician suggests having his wife come to clinic with her passport in order to receive additional ARVs to make up for those shared, client agrees to set up reminder calls with his clinic so he is not in the scenario of running out of ARVs and needing to use someone else's, and client decides to use a calendar to record upcoming appointment dates. |
| Sickness Related Issues | 2 | 0 | Client missed their appointment because they were feeling sick and came to clinic once they felt better → Clinician states that, in this scenario, the client did the right thing to not send a guardian because the client also needed to get a blood draw. Clinician affirms that client should do the same thing next time (simply come to clinic as soon as he feels ok again) so long as they have enough medication around. |
| Trouble Remembering Appointments & Medication Schedule | 5 | 1 | Client states they are having difficulty remembering to take their medications → Clinician and client brainstorm ideas for remembering and client decides to post a timetable on their wall to remind them. |
| Conversations about Lab Tests | 1 | 0 | Client did not understand what their lab tests meant and thus did not come to clinic → Clinician walks the client through how to read and interpret the HIV lab tests and explains that clients should ask someone at the facility, upon receipt of the results, to aid in test interpretation if there is any confusion. |
| Issues Accessing HIV-Related Services | 2 | 1 | Client had issues navigating HIV services → Clinician offers to escort the client in real time to pick up the client's medications so that the client will feel comfortable going forward. |
| Familial / Personal Issues | 1 | 0 | Client explains that, upon initial receipt of his diagnosis, he was not mentally or emotionally prepared to undergo treatment. He is currently experiencing marital strife, though, and for fear of the fate of his children, he wants to start treatment → Clinician reexplains to the client how to take his medications properly and counsels on what to do if the client must leave town. |
| Transportation | 2 | 0 | Client explains that they did not come to their recent appointment because they ran out of money → Clinician and client brainstorm and decide that the client should set aside some money each month which can be used for transport. When their appointment is nearing and they find that the money is still available, they should come to the hospital while the money is available even if it means coming earlier than the set appointment date. |

